## Supplementary Methods for "Integrated methylome and phenome study of the circulating proteome reveals markers pertinent to brain health"

**SOMAscan measurement quality control**

The following information outlines each stage of the quality control processes carried out within the SOMAscan® V4 platform ^1^. Of the 5,284 reagents, there are 12 are spike-in controls, 286 negative control/non-human targets, 7 deprecated and 4,979 human SOMAmers that target 4,776 unique proteins. These 4,979 SOMAmers are spread across three dilution bins as follows: 160 in the 0.005% bin, 797 in the 0.5% dilution group and 4,022 reagents in the 20% bin. Regarding the 96-well plates, 11 wells are replicate controls (5 calibrator samples, 3 quality control samples and 3 buffer or no protein samples) and 85 are for biological samples.

- **Hybridisation control normalisation** is applied to control for nuisance variance within individual wells. A scaling factor is calculated as the median ratio of reference relative fluorescence intensities (RFUs) for 12 spike-ins against the observed RFUs in that sample or well. The reference RFUs are the median RFUs of these control SOMAmers across the entire plate of samples.
- **Intra-plate median signal normalisation** is performed to minimise variation across wells in a plate that might be caused by variability in pipetting, reagent concentration, washing steps, assay timing and differences in overall input protein concentration. This is applied separately to wells of the same class (i.e. separately for each buffer, calibrator, quality control type) and within SOMAmers of the same dilution factor (0.005%, 0.5%, and 20%). This creates a number of sample-SOMAmer groupings. The RFU of each SOMAmer (within a sample-SOMAmer group) is divided by the median of this SOMAmer’s RFUs across the entire plate. Then, a scale factor is applied to each well but only for SOMAmers in the SOMAmer-sample grouping. The scale factor associated with a given well is calculated as the inverse of the median ratio for that sample across all SOMAmers in the sample-SOMAmer grouping. In a given sample, RFUs for the SOMAmer in this grouping are median-normalised by multiplying RFUs by the scaling factor.
- **Calibration Normalisation** accounts for variability across plates within a run. This is typically caused by variability introduced by differences in scanner intensity. RFUs for dedicated calibrator samples in a plate are each divided by a reference value. The median of this ratio across calibrators in a plate is used to calculate a single scaling factor for the plate.
- **Calibration** refers to a normalisation procedure that accounts for variability between assay runs and/or experiments. This is performed on a SOMAmer-by-SOMAmer basis. Dedicated calibrator controls are utilised in this step. A SOMAmer-specific reference value is divided by the median of calibrator control RFUs and this gives the calibration scaling factor for the SOMAmer across the entire run.
- **Adaptive Normalisation by Maximum Likelihood** is an optional step which was performed in the Generation Scotland cohort that utilises estimates for the median signal and median absolute deviation of each SOMAmer taken from a reference sample (n ~ 1,000). This is performed separately for each dilution bin. This method provides a scaling factor for the SOMAmer that maximises the probability that a sample’s RFU comes from the sampling distribution. The method assumes that more than 30% of analytes are consistent with reference-based assumptions. Adaptive normalisation reduces technical variability between wells and inter-sample biological variability contributing to differences in total protein signal.
- **Post-calibration quality control** is carried out after the above steps. Three pooled quality control replicates are randomly distributed on the 96-well plate. For each SOMAmer, the accuracy of the median replicate signal on the plate is compared against a reference value. The result is a vector of quality control accuracy ratios across the SOMAmers. This provides information on whether there is still significant post-calibration variability and also on the quality of each assay run. In total, at least 85% of quality control ratios must be between 0.8 and 1.2 in a plate to meet acceptance criteria. Furthermore, a plate also has to show plate scaling factors between 0.4 and 2.5 prior to acceptance and release.

**DNAm measurement and quality control**

DNA methylation in the STRADL subset of Generation Scotland was assayed into two distinct sets (n_set1_=504, n_set2_=306). *Meffil* was used to exclude individuals with a mismatch between DNAm-predicted sex and recorded sex, samples in which greater than 0.5% of CpGs had a detection *P*>0.01, outliers for bisulphite conversion control probes, samples with median signal intensity >3 standard deviations lower than expected and samples with evidence of dye bias ^2^. *shinyMethyl* was then used to exclude outliers based on visual inspection of plots with the log median intensity of methylated versus unmethylated signals per array ^3^. Following this, *Meffil* was used to identify and exclude probes that had a beadcount of less than 3 in greater than 5% of samples and/or probes for which >1% of samples had a detection *P*>0.01. Plots from multidimensional scaling (MDS) were investigated to inspect for further outlier samples. Forty males were identified as outliers according to X chromosome DNAm levels. These participants were removed from the analyses. Data were re-normalised and inspection of MDS plots confirmed that no further outliers were present across both sets. Data were then normalised using the dasen method in *wateRmelon* ^4^ and converted to M values using the beta2m function in *lumi* ^5^*.* There were a total of 778 individuals with methylation and proteomic data available.

**Cognitive scores**

The sum of immediate and delayed recall of one oral story from the Wechsler Logical Memory Test was taken as the logical memory phenotype (maximum score of 25 for each recall test with a combined maximum score of 50) ^6^. Details about the stories that were remembered correctly were recorded as individual points contributing to the scores. The Wechsler Digit Symbol Substitution Test, which requires individuals to recode digits to symbols and represents a count of correct pairs within a timeframe of 120 seconds was used to measure processing speed phenotype ^7^. The verbal reasoning phenotype measures verbal comprehension and phonemic fluency and was based on the Controlled Oral Word Association task ^8^ with letters C, F and L. The number of words named starting with the given letters in a one minute period were recorded as the score. The Mill Hill Vocabulary test was used as a measure of acquired verbal intelligence, and is an estimate of ‘crystallised intelligence’ and peak cognitive ability. This records the number of times participants successfully explain the meaning of words select a synonym, using junior and senior synonyms ^9^. The Matrix Reasoning test, a paper adaptation of the computerised version from the COGNITO psychometric examination ^10^ was used to measure perceptual organisation and visuospatial logic. The Matrix Reasoning test measures non-verbal, abstract reasoning and records the number of correct answers recognising the missing element in a pattern that is presented as a matrix. Outliers were defined as scores >3.5 standard deviations above or below the mean and were removed prior to analysis.

The first unrotated principal component combining logical memory, verbal fluency, vocabulary and digit symbol tests (sum of squares loadings = 1.76 and proportion of variance = 0.44) was calculated as a measure of general cognitive ability (‘*g’*). General fluid cognitive ability (‘*gf*’) was extracted using the same approach, but with the vocabulary test (a crystallised measure of intelligence) excluded from the composite (sum of squares loadings = 1.52 and proportion of variance = 0.51). While highly similar to *g*, the *gf* score is exclusive to measures such as memory and processing capability that are considered fluid. *gf* may therefore be of pertinence to delineate when assessing cognitive performance in ageing individuals. Maximum possible scores have been summarised in the main body of the manuscript and can also be found in Habota *et al*, 2019 ^11^.

**Brian imaging protocol**

The STRADL subset of Generation Scotland included participants that were scanned at two centres: the Ninewells Hospital in Dundee and the Aberdeen Royal Infirmary in Aberdeen. Participants in Dundee were scanned using a Siemens 3T Prisma-FIT (Siemens Healthineers, Erlangen, Germany) with a 20-channel head and neck coil and back-facing mirror (software version VE11, gradient with max amplitude 80 mT/m and maximum slew rate 200 T/m/s). Participants in Aberdeen, were imaged using a 3 T Philips Achieva TX series MRI system (Philips Healthcare, Best, Netherlands) with a 32-channel phased-array head coil and back-facing mirror (software version 5.1.7; gradients with maximum amplitude 80 mT/m and maximum slew rate 100. Both centres followed the same protocol including structural sequences ^11^. 3T MRI scans were anonymised at the time of acquisition. The structural sequences were as follows: 3D T1-weighted fast gradient echo with magnetisation preparation; 3D T2-weighted fast spin echo; 3D Fluid Attenuation Inversion Recovery (FLAIR); Diffusion Tensor Imaging (DTI); Susceptibility Weighted Imaging (SWI) or T2*-weighted gradient echo ^11^.

**Volumetric brain imaging**

To generate volumetric data, scans were processed using FreeSurfer version 5.3 ([http://surfer.nmr.mgh.harvard.edu](http://surfer.nmr.mgh.harvard.edu/), RRID:SCR_001847 ^12^). Any major errors in segmentation or cortical parcellation were excluded from analyses based on visual inspection. Manual brain mask edits were made when the skull was included in parcellations to remove the skull from parcellated tissue. If parts of the brain were not included in the parcellation manual edits were made to delineate the boundaries to include missing tissue. A record of edits was kept and participants were given a score of 0 if they had no manual edits to their scan or a score of 1 if they had either an edit. This was used as a covariate in statistical analyses since the process of QC and manual editing of scans is a subjective process which could introduce bias. To derive white matter hyperintensity volumes, FLAIR (and T1) scans were processed using the Lesion Segmentation Toolbox (LST) version 3.0.0 for SPM ([www.statisticalmodelling.de/lst.html](http://www.statisticalmodelling.de/lst.html)). Within the LST the white-matter lesions are segmented with the lesion growth algorithm ^13^. The algorithm first segments the T1 images into the three main tissue classes (cerebrospinal fluid, grey matter and white matter) and this information is combined with FLAIR intensities to calculate lesion belief maps. These maps are thresholded with a pre-chosen initial threshold (κ parameter) to obtain an initial binary lesion map. This map is then grown along voxels that appear hyperintense in the FLAIR image, which produces a lesion probability map. The lesion probability map is then again thresholded to define WMH regions. Volumetric measurements of whole brain volume (no ventricles), global grey matter volume and white matter hyperintensity volume were used structural indicators of brain health in present study. Intracranial volume was also available as a covariate to adjust for head size in all volumetric analyses.

**Fazekas white matter hyperintensity scores**

Severity of white matter hyperintensities were graded according to the Fazekas scale ^14^ that distinguishes periventricular and deep white matter hyperintensities and grades them from 0 (absent) to 3 (severe). A score of 1 is defined as caps or pencil thin lining or punctuate foci; a score of 2 is defined as a smooth halo or beginning to confluence and a score of 3 is defined as irregular periventricular signal extending into the deep white matter or large confluent areas. While Fazekas scores are considered less precise than direct measurements of white matter hyperintensity volumes, they were included for comparison in the present study.

**Diffusion-weighted imaging**

Diffusion-weighted imaging was performed at the same two sites and with the same scanners as T1-weighted imaging, as part of a single protocol. For DTI data, pre-processing and quality control was performed using tools available from FSL (<https://fsl.fmrib.ox.ac.uk/fsl/fslwiki>). Briefly this included (1) correcting for eddy current-induced distortions and subject movement while in scanner; (2) skull stripping using BET at a threshold of 0.2; (3) using DTIFIT in order to compute diffusion tensor characteristics (i.e. principal eigenvectors or V1, V2, V3; eigenvalues or L1, L2, L3; fractional anisotropy (FA), mean diffusivity (MD); and (4) visually checking the quality of FA images at this stage in order to exclude any distorted images. There were three tracts not included in any tract categories: corpus callosum, corona radiata and internal capsule. The five unilateral tracts included the corpus callosum, fornix and the body, genu and splenium of the corpus callosum. Tract Based Spatial Statistics (TBSS) was carried out according to the ‘The Enhancing NeuroImaging Genetics through Meta-Analysis’ (ENIGMA) Consortium DTI protocol (<http://enigma.ini.usc.edu/protocols/dti-protocols/>). Region of interest (ROI) extraction analyses were then performed also using ENIGMA protocols to extract [fractional anisotropy](https://www.sciencedirect.com/topics/neuroscience/fractional-anisotropy) (FA) and [mean diffusivity](https://www.sciencedirect.com/topics/immunology-and-microbiology/mean-diffusivity) (MD) measures (<http://enigma.ini.usc.edu/protocols/dti-protocols/>). White matter tracts were categorised using the Johns-Hopkins University DTI-based white matter atlas ^15^. This resulted in 5 unilateral tracts and 19 bilateral tracts, as well as an average measure for FA and MD. The general FA and MD measures were used in the present study.

**Characterisation of *cis* and *trans* pQTMs**

pQTMs were categorised into *cis* and *trans* effects. *Cis* pQTMs were defined as having CpG sites that were within 10 Mb of the transcription start site (TSS) of the gene encoding the target protein of interest, while also being situated on the same chromosome as the protein-coding gene. *Trans* pQTMs were defined as having CpGs that lied outside of this 10Mb region surrounding the TSS of the gene, or were located on a chromosome distinct from the chromosome on which the protein target’s TSS was located. TSS positions were catalogued using *biomaRt* and Ensembl v83 ^16,17^.
