## Supplementary Figures for "Integrated methylome and phenome study of the circulating proteome reveals markers pertinent to brain health"

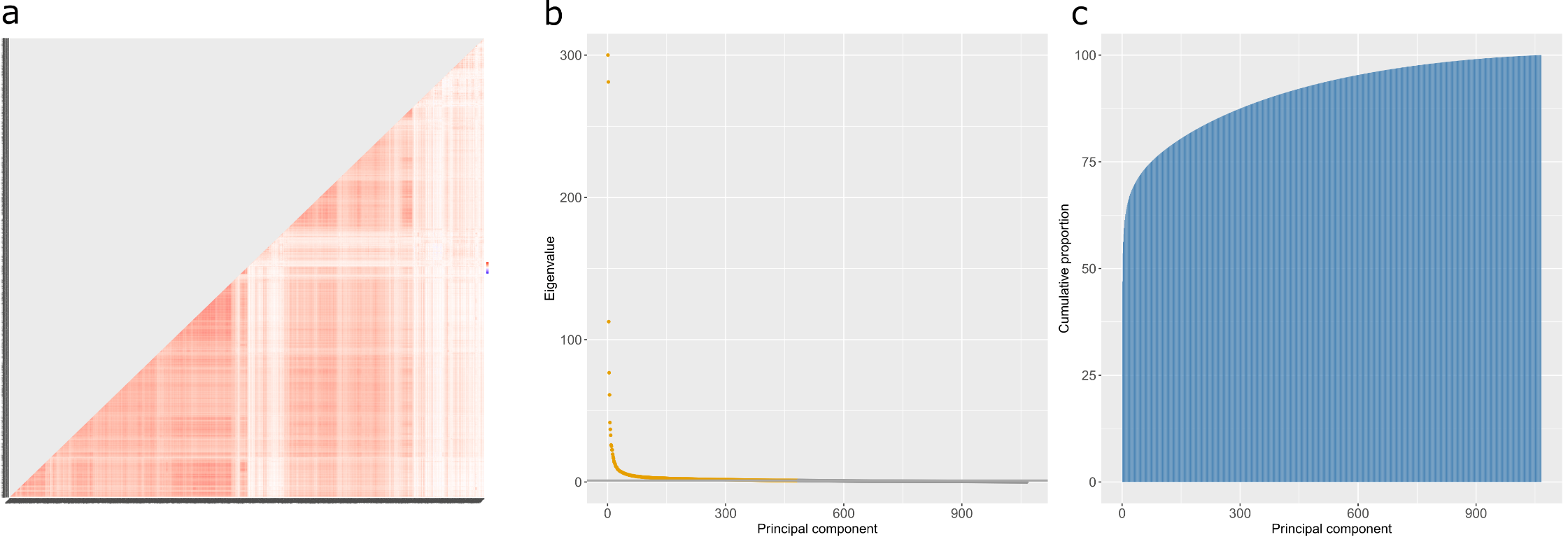

**Supplementary Figure 1. Correlation heatmap and principal components analyses for 4,235 SOMAmer measurements (corresponding to 4,058 unique protein levels) included in MWAS and PheWAS analyses. a** Correlation structures are shown for the 4,235 SOMAmer measurements. Eigenvalue coefficients **b** and the cumulative proportion of variance explained across each of the principal components **(c)** are presented for the 4,235 SOMAmer measurements; 483 eigenvalues > 1 are denoted in yellow. 143 components explained at least 80% of the variance in protein levels. The eigenvalue for the first component was 1986, however this has been set to 300 for the purposes of this visualisation. A full numeric summary of cumulative variance and respective eigenvalues for components is provided in Supplementary Table 3.

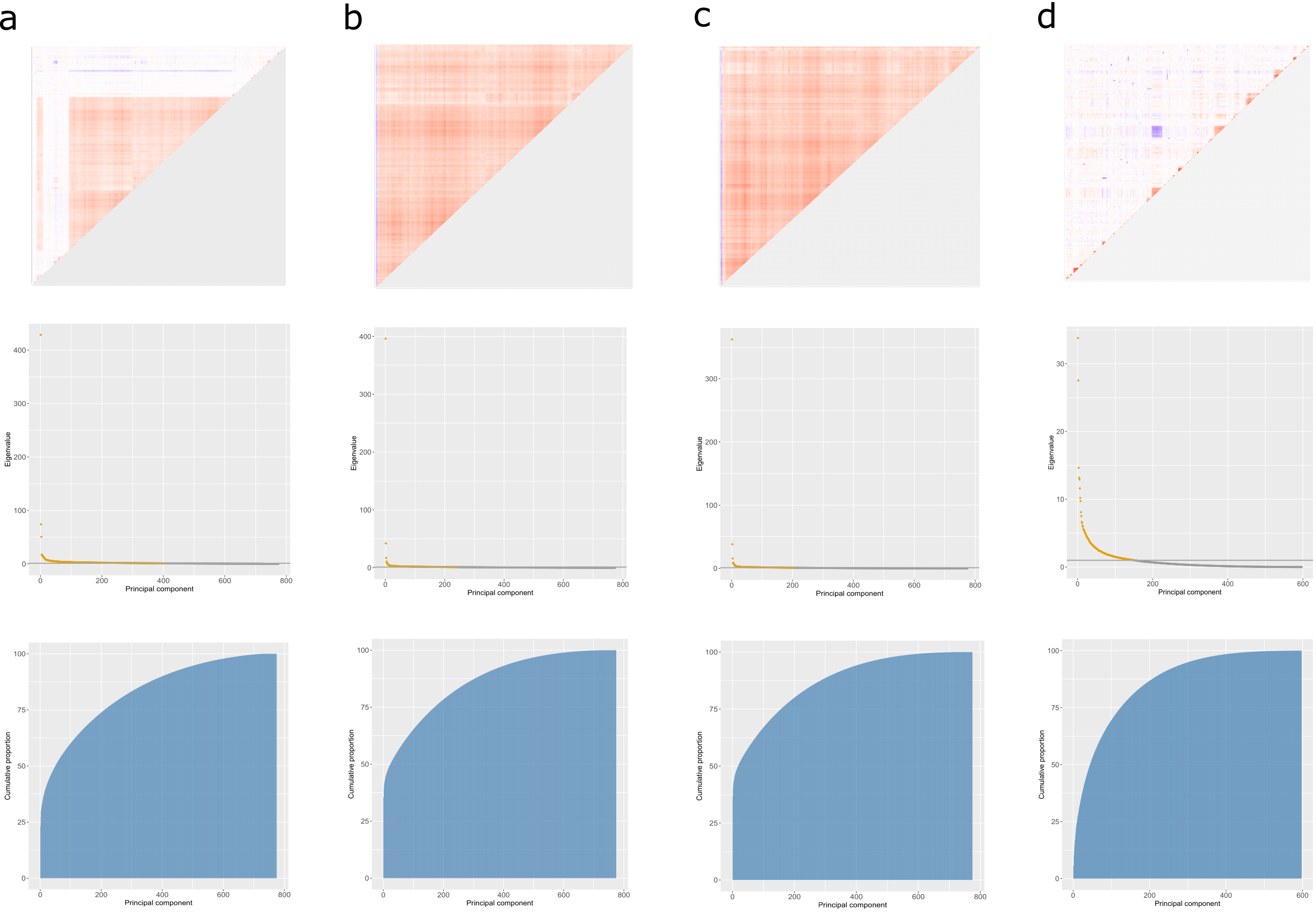

**Supplementary Figure 2. Correlation structure heatmaps (upper row), eigenvalues (middle row) and cumulative variance (lower row) from principal components analysis plots for CpGs involved in fully-adjusted MWAS pQTMs.** **a** All of the 1,837 unique CpGs that were involved in 2,928 total associations. **b** The 1,116 CpGs that were associated with PRG3 levels. **c** The 987 CpGs that were associated with PAPPA levels. **d** The 597 CpGs that were involved in the 825 pQTM associations that did not include PRG3 or PAPPA protein levels in the fully-adjusted MWAS.

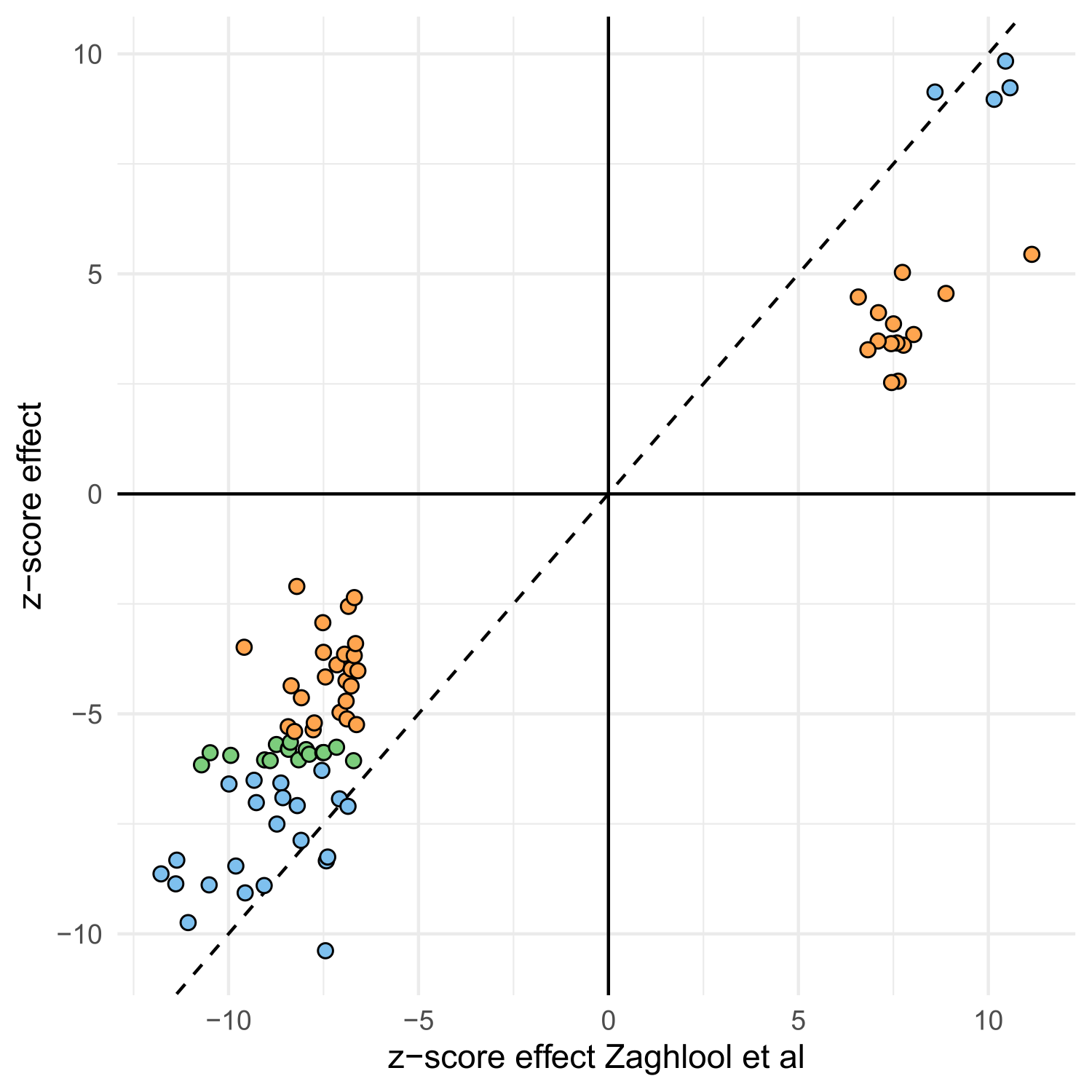

**Supplementary Figure 3. Effect coefficients (in z-score stadardised format) plotted for 81 comparable pQTM associations between Zaghlool et al and the present MWAS.** 26 associations that replicated with our significance threshold for the fully-adjusted MWAS (4.5x10^-11^) are shown in blue, whereas 16 further associations that replicated at the standard MWAS threshold of significance (3.6x10^-8^) are shown in green. Orange points indicate the 39 remaining associations that replicated at Nominal P < 0.05. A full numeric summary of these data is provided in Supplementary Table 11.

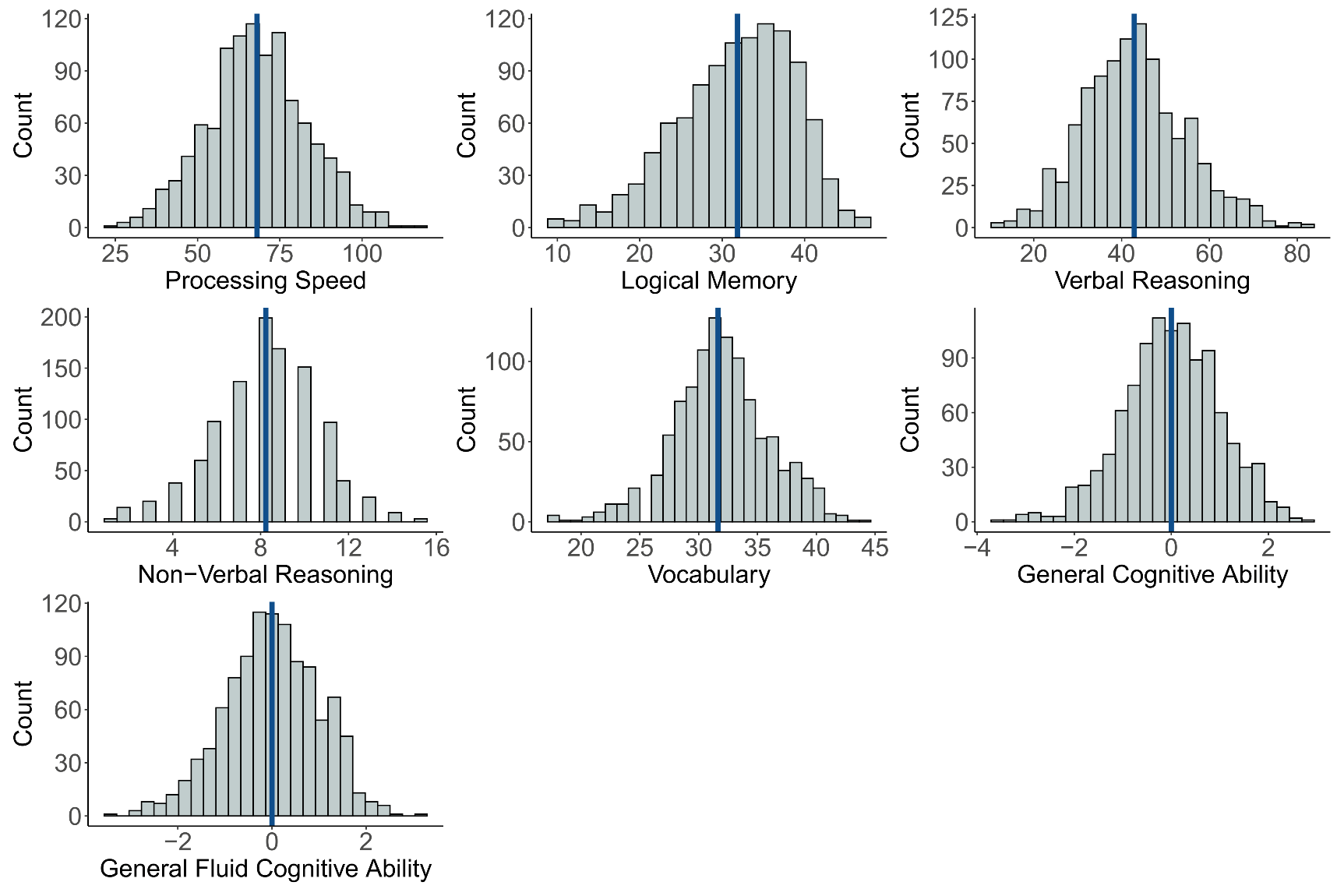

**Supplementary Fig. 4. Histogram distribution plots for cognitive scores in the Generation Scotland sample used in the protein PheWAS.** There were 1,053, 1,060, 1,058, 1,060, 1,058, 1,049 and 1,051 individuals with Processing Speed, Logical Memory, Verbal Reasoning, Non-Verbal Reasoning, Vocabulary, General Cognitive Ability and General Fluid Cognitive Ability measures included in lmekin association models, after accounting for 5 individuals with missing depression status data. The mean of each score is annotated as a vertical blue line. All variables were trimmed to remove outliers (>3.5 SD from mean) before plotting.

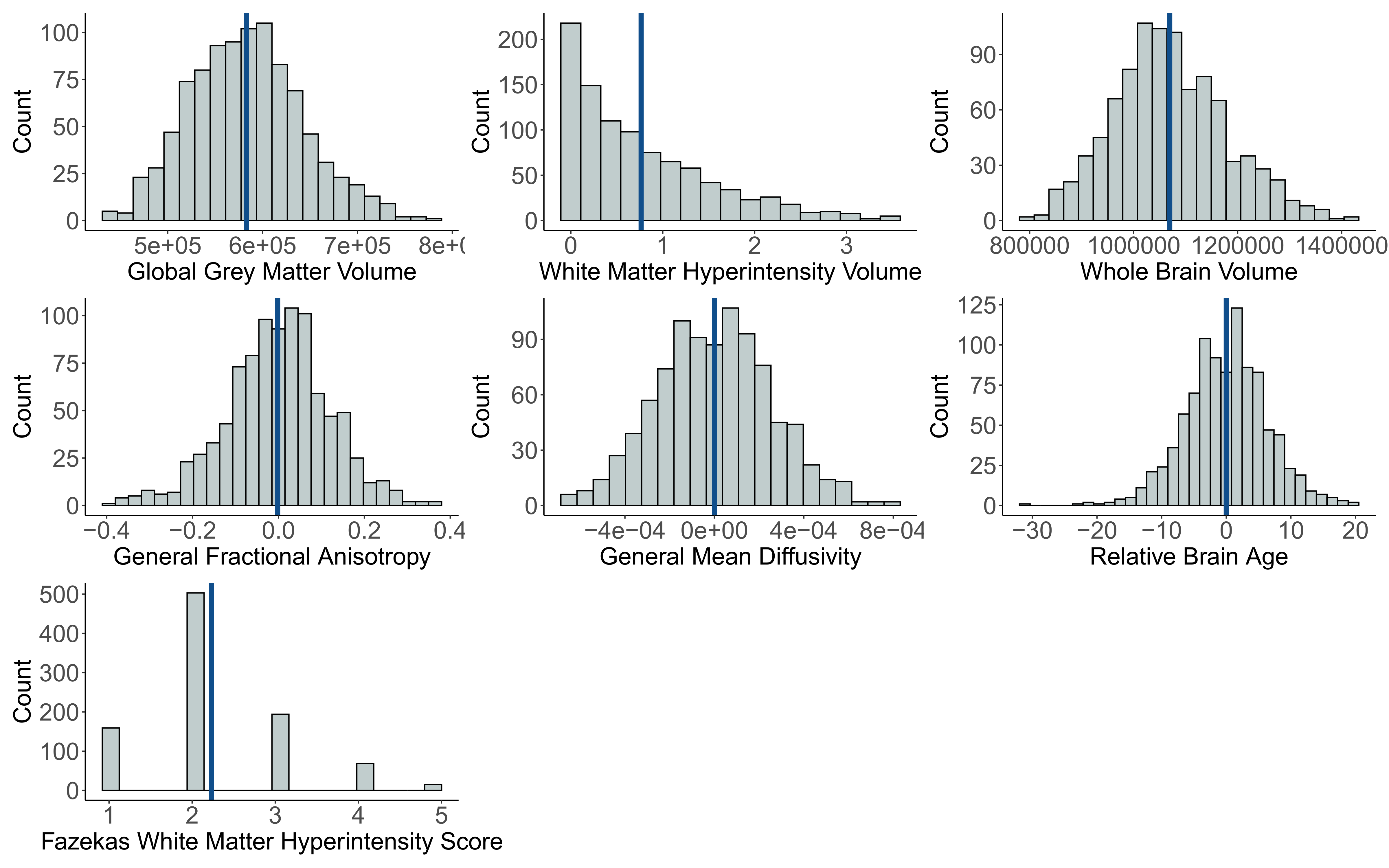

**Supplementary Fig. 5. Histogram distribution plots for brain imaging phenotypes in the Generation Scotland sample used in the protein PheWAS.** There were 940, 927, 939, 921, 920, 937 and 963 individuals with Global Grey Matter Volume, White Matter Hyperintensity Volume, Whole Brain Volume, General Fractional Anisotropy, General Mean Diffusivity, Fazekas White Metter Hyperintensity Scores and Relative Brain Age measures included in lmekin association models, after accounting for 5 individuals with missing depression status data. The mean of each score is annotated as a vertical blue line. All variables were trimmed to remove outliers (>3.5 SD from mean) before plotting. A log + 1 transformation was applied to White Matter Hyperintensity Volume.

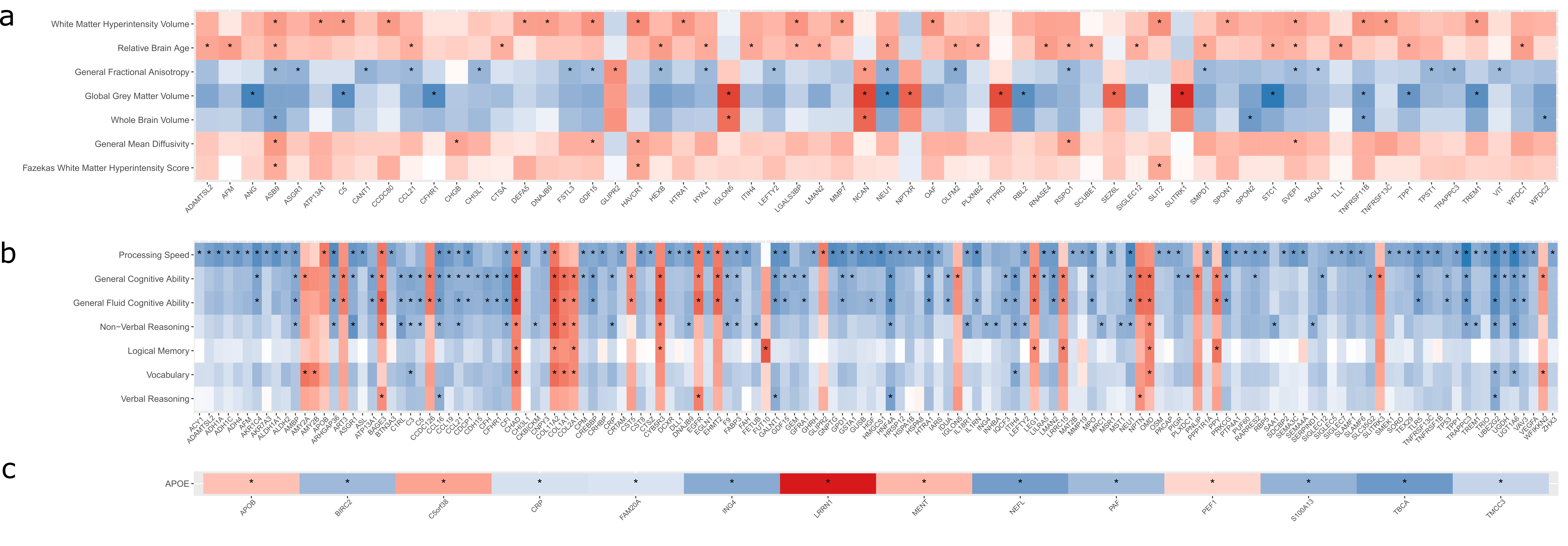

**Supplementary Fig. 6.** **Standardised beta coefficients plotted from phenome-wide protein association studies (protein PheWAS) between 4,058 protein levels and 15 neurologically-relevant phenotypes in Generation Scotland (maximum N=1,095).** All 405 associations between plasma protein levels and either brain imaging **(a)** cognitive scoring **(b)** or APOE haplotype **(c)** that had P < 3.5x10^-4^ are shown in this heatmap and are indicated by an asterisk. 14 *APOE* haplotype (involving 14 unique proteins), 296 cognitive scoring (involving 142 unique proteins) and 95 brain imaging (involving 60 unique proteins) associations are presented. Negative and positive direction of effects are shown in blue and red, respectively. There were 191 unique protein levels involved in the 405 associations; these associations are presented in Supplementary Table 17.

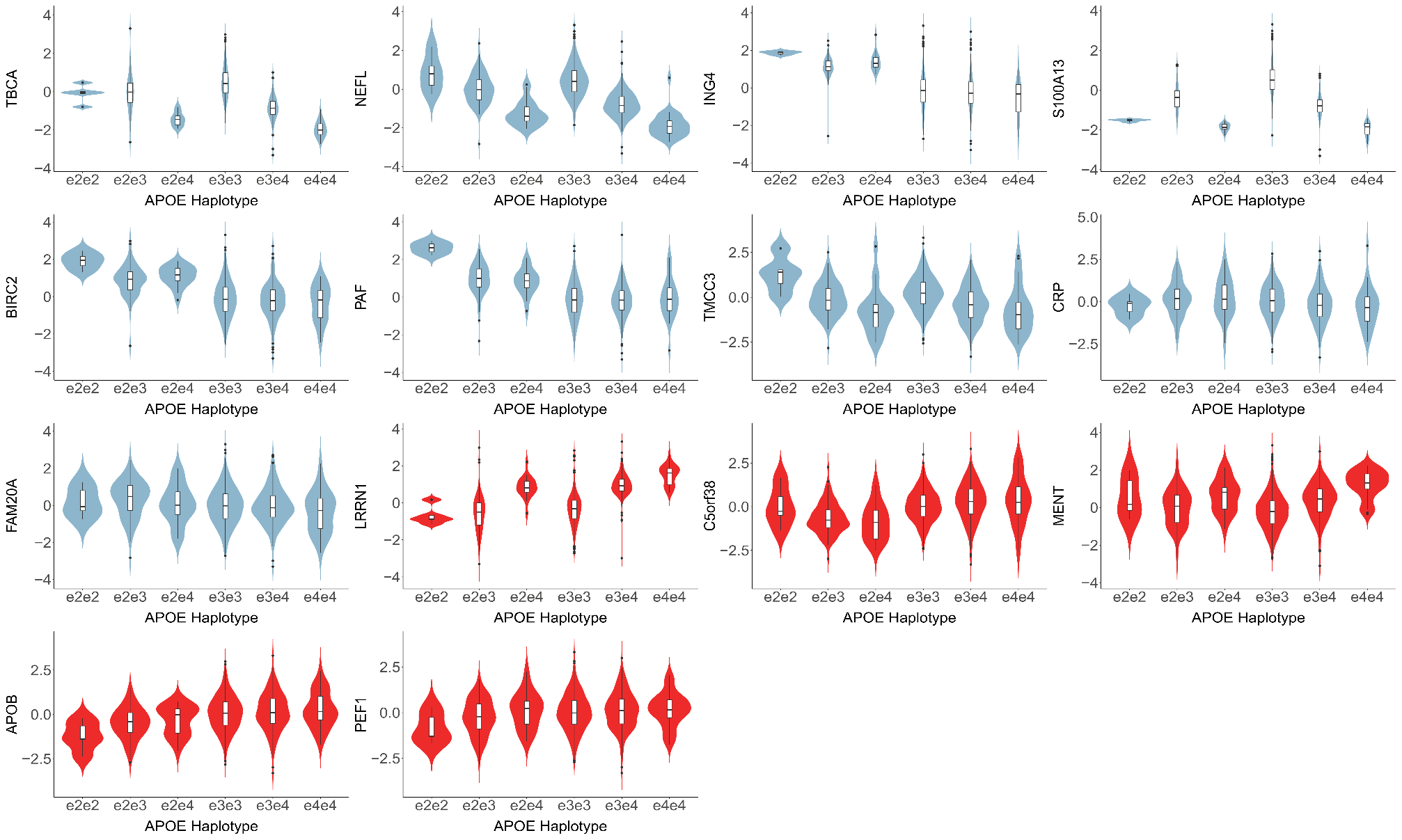

**Supplementary Fig. 7. Violin boxplots for associations between the levels of 14 proteins and *APOE* haplotype that had P < 3.5x10^-4^ in the protein PheWAS (N e2e2=5, N e2e3 = 121, N e2e4=22, N e3e3=633, N e3e4=234, N e4e4=35).** Associations with positive beta coefficients (red) indicate that increased levels of the protein associate with the presence of one or two copies of the *APOE* e4 allele. Associations with negative beta coefficients (blue) indicate inverse relationships between increased levels of the protein and one or two copies of the *APOE* e4 allele. A total of 1023 individuals were included in the analyses (1,065 individuals, excluding 15 missing *APOE* status, 5 missing depression status and 22 individuals with e2e4 that were not part of the e2/e3/e4 groups used in the analysis. These associations can be found in Supplementary Table 15.

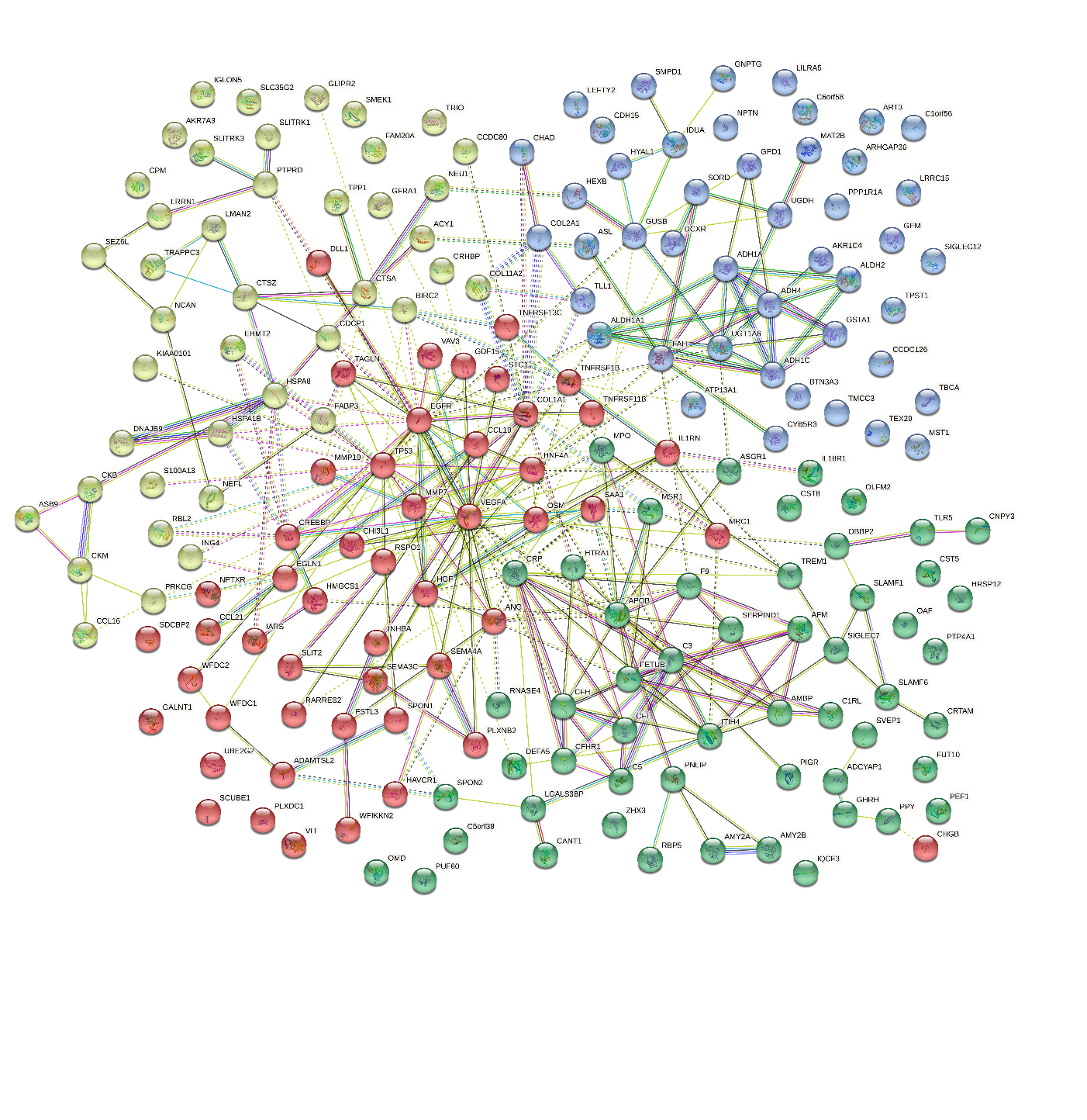

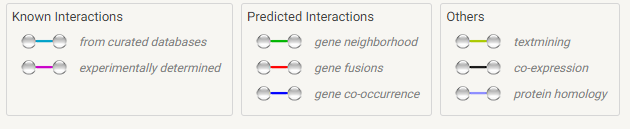

**Supplementary Figure 8.** **STRING protein interaction network created based on the corresponding genes for the 191 proteins that had P<3.5x10^-4^ in associations with either brain imaging, cognitive scoring or *APOE* haplotype.** Proteins are shown as nodes in the network, with connecting edges representing projected commonality in function between protein pairs. An index describing the source of information that was used to inform edge connections is included. BAGE3 did not have a gene name entry in the STRING database and is therefore excluded from this plot.

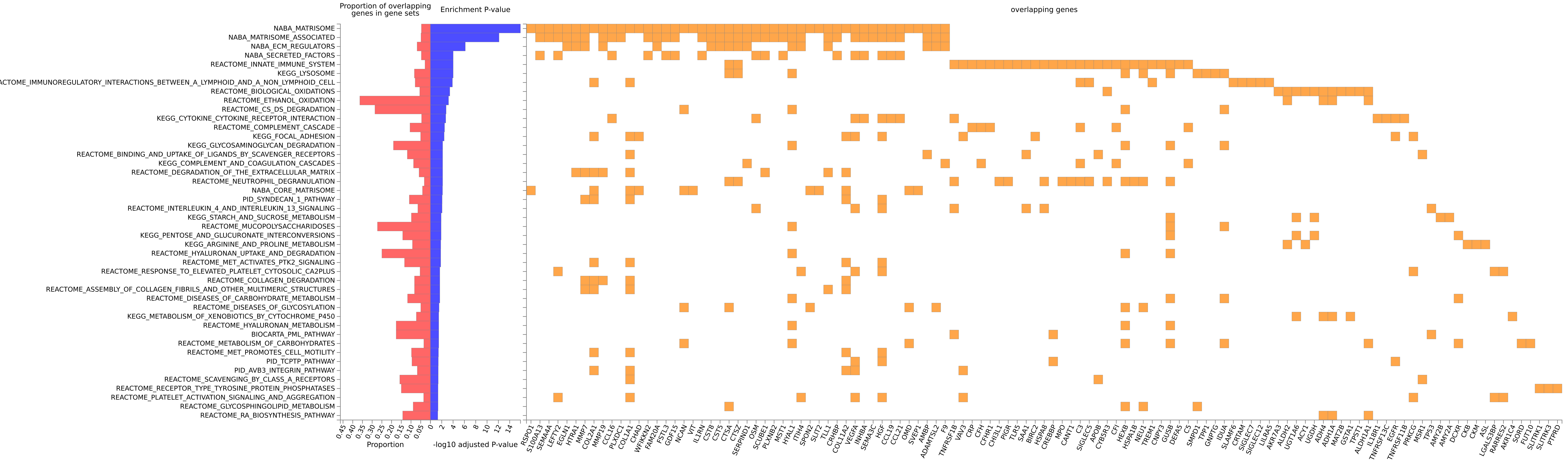

**Supplementary Figure 9. FUMA gene-set enrichment for the genes corresponding to the 191 proteins that had P<3.5x10^-4^ in associations with cognitive scoring, brain imaging or *APOE* haplotype in the protein PheWAS.** The proportion of overlapping genes for each set is described, with the estimated P-value for enrichment plotted on a log10 scale. All gene sets shown had FDR-adjusted P < 0.05 and a minimum of 3 overlapping genes.

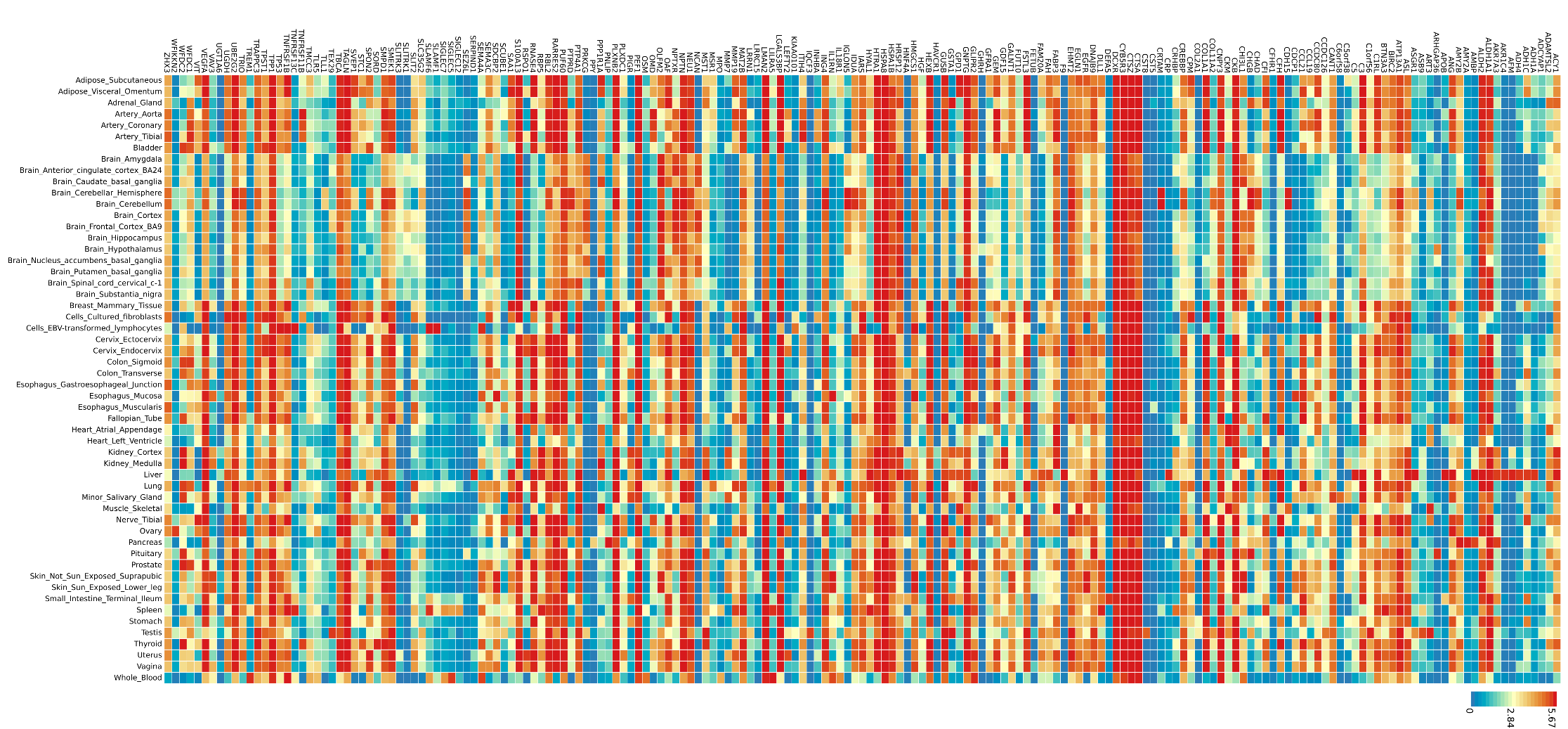

**Supplementary Figure 10. FUMA tissue expression heatmap for the 191 genes that were associated with either brain imaging, cognitive scoring or *APOE* haplotype in the protein PheWAS.** Average expression is provided in log2 transformed scale. Red rectangles indicate higher expression, whereas blue rectangles indicate lower expression. Genes are ordered alphabetically.

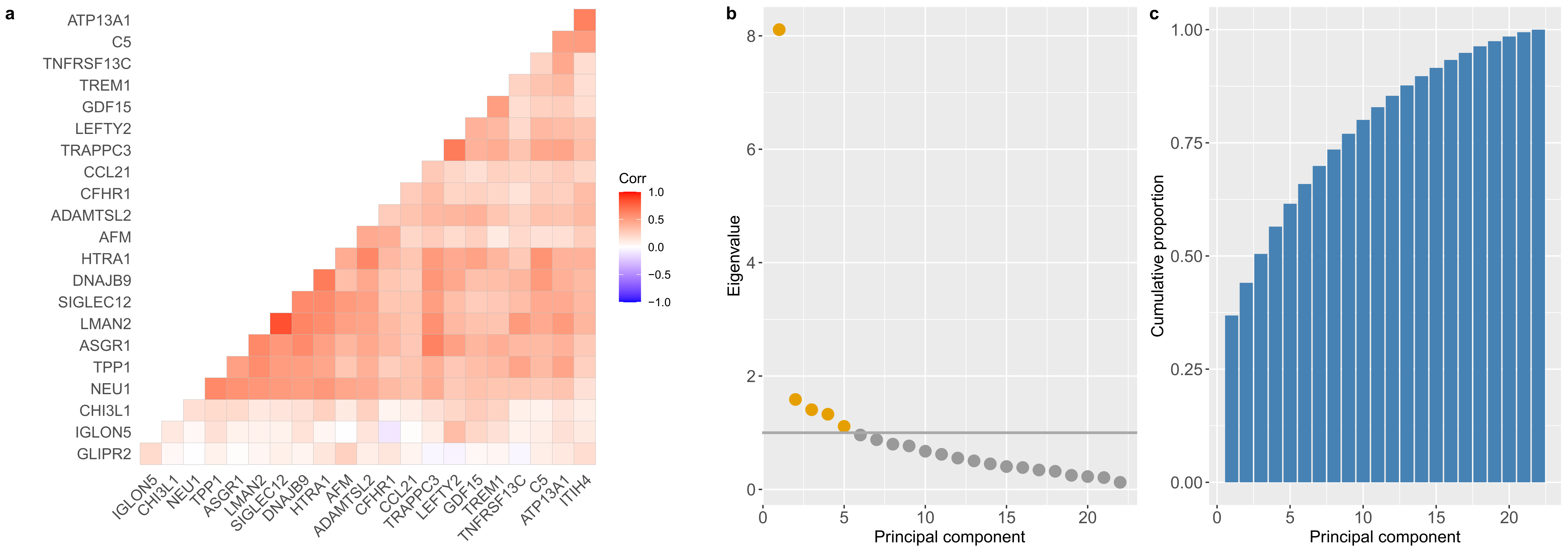

**Supplementary Figure 11. Correlation heatmaps and principal components analyses for 22 protein levels that had P<3.5x10^-4^ in associations with both a brain imaging and a cognitive scoring measure. a** Correlation structures are shown for the 22 protein levels. Eigenvalue coefficients **b** and the cumulative proportion of variance explained across each of the principal components **c** are presented for the 22 proteins; eigenvalues > 1 are denoted in yellow.

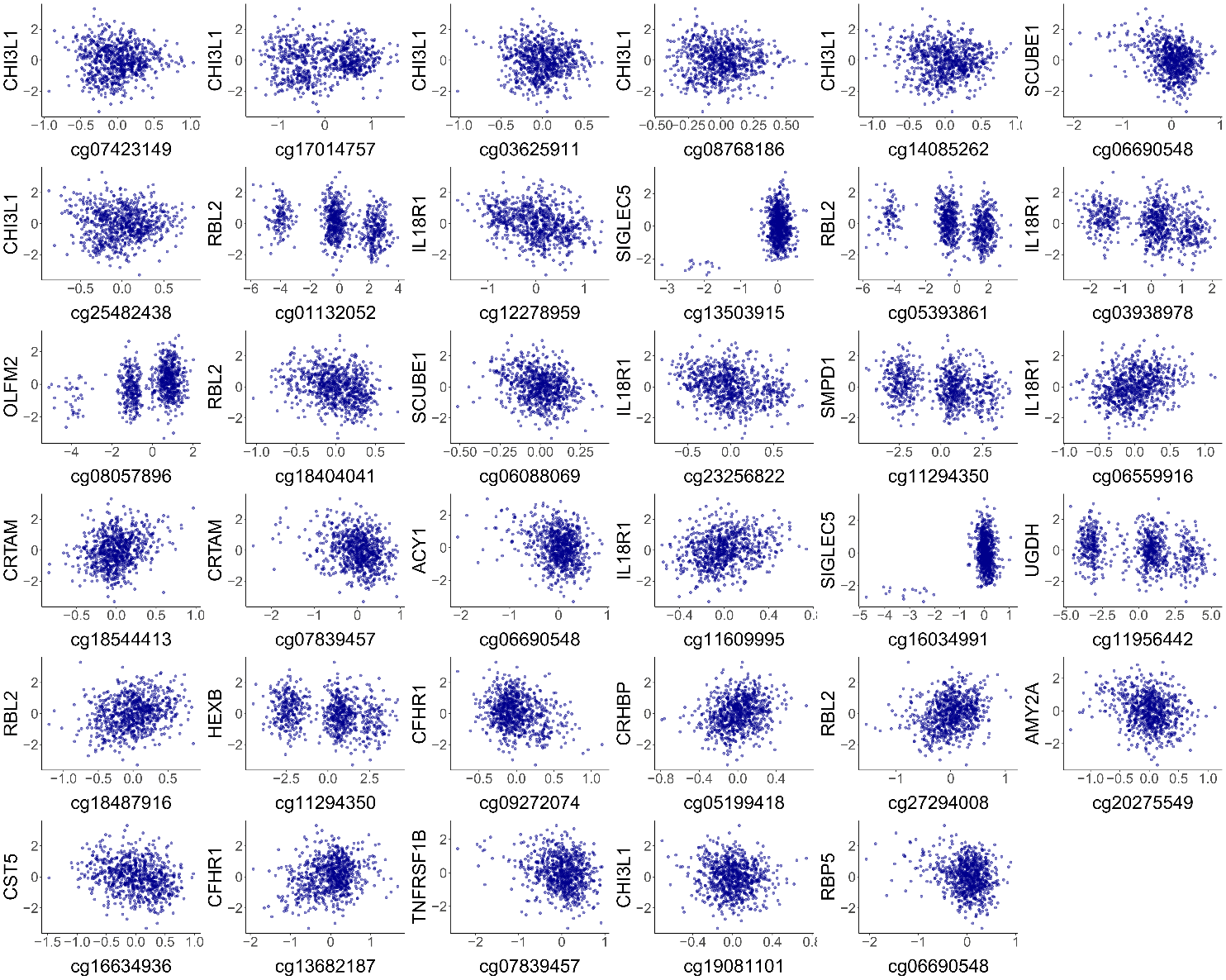

**Supplementary Figure 12. Association plots for the 35 pQTMs involving protein makers of brain health outcomes (N=744).** DNAm at each CpG is plotted on the x-axis, against the levels of each protein on the y-axis. DNAm and protein data from the fully-adjusted MWAS were used in all cases. Trimodal CpG distributions indicate the presence of a likely underlying mQTL genetic effect that may partially contribute to DNAm at the CpG site.

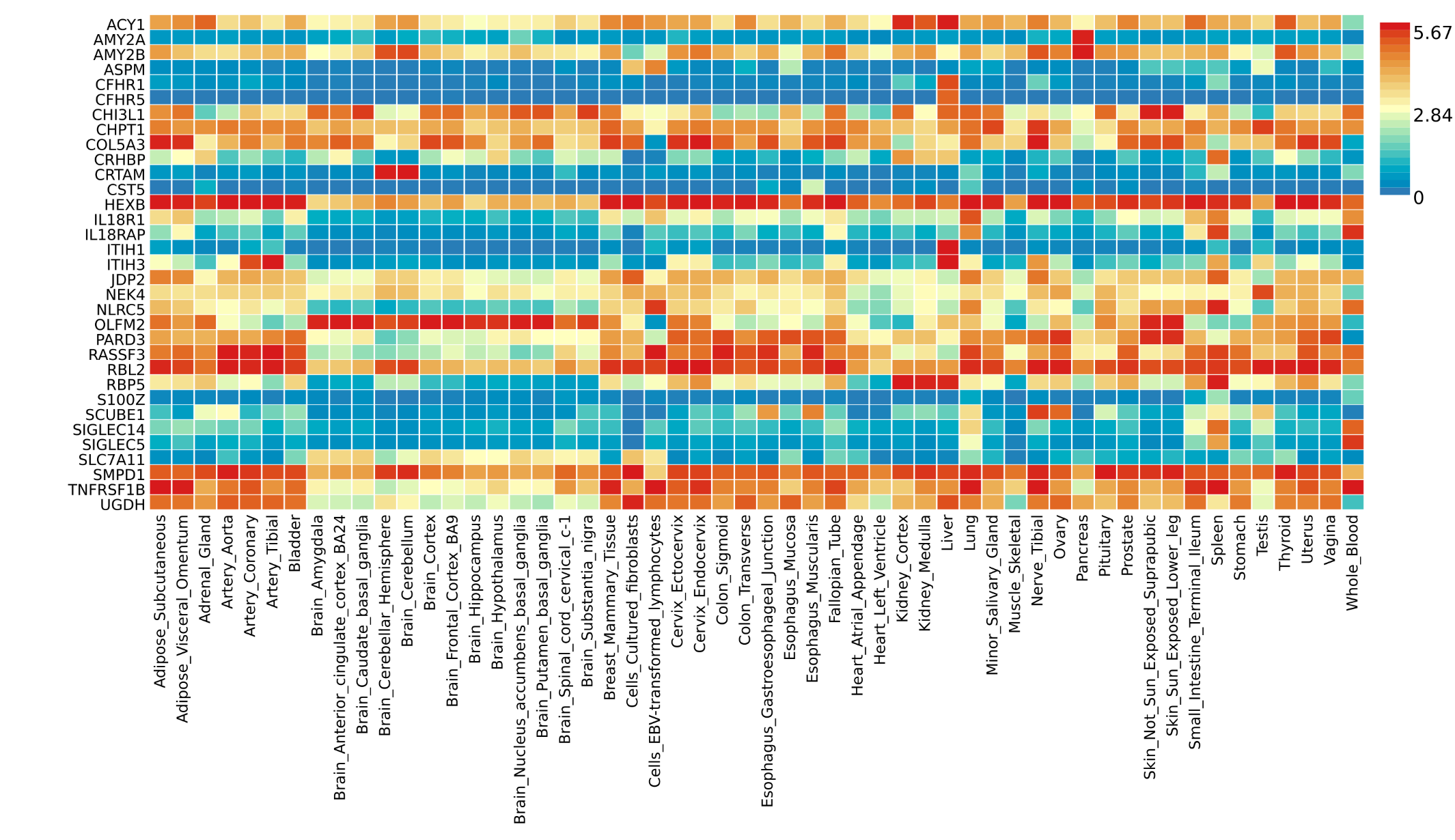
**Supplementary Figure 13. FUMA tissue expression heatmap for the 33 genes that were either implicated by proteins or CpGs in the 35 neurological pQTMs.** Average expression is provided in log2 transformed scale. Red rectangles indicate higher expression, whereas blue rectangles indicate lower expression. Genes are ordered alphabetically.

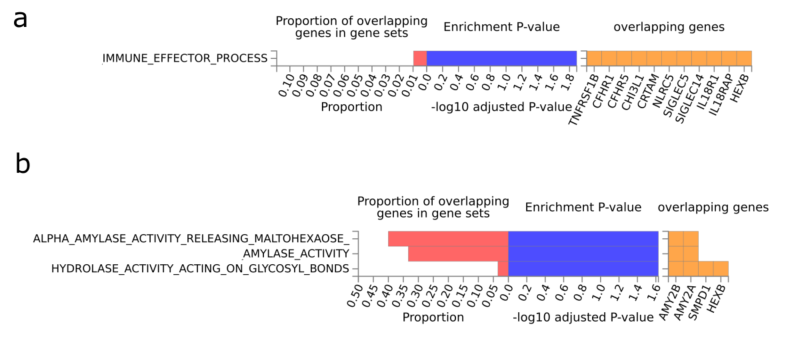

**Supplementary Figure 14. FUMA gene-set enrichment for the 33 genes corresponding to either CpG or protein levels in the subset of 35 neurological pQTMs from the protein MWAS.** The proportion of overlapping genes for canonical pathways (**a**) and biological pathways (**b**) is described, with the estimated P-value for enrichment plotted on a log10 scale. All gene sets shown had FDR-adjusted P < 0.05.

**Supplementary Figures 15-21. CHIP-seq and promoter-centered Hi-C chromatin mapping plots between genes harboring CpGs and the respective protein-coding genes for all cis pQTMs that had loci within the same chromosomes.** Seven plots are listed in the following order: S100Z-CRHBP, SIGLEC14-SIGLEC5, IL18RAP-IL18R1, COL5A3-OLFM2, CFHR5-CFHR1, ASPM-CFHR1 and AMY2B-AMY2A, where the first gene represents the CpG gene and the second the protein-coding gene of interest. Genomic positioning is indicated, with grey vertical lines demarcating the CpG position. CHIP-seq intensities are shown for activating marks H3K4me1 and H3K27ac. CHIP-seq intensities for the silencing marks H3K27me3 (that can also be associated with promoter regions or bivalent chromatin) and H3K4me3 (which is generally a bivalent mark when flanking active chromatin) are also shown. All marks are shown for peripheral bloody mononuclear cells (PBMCs) and brain hippocampus. Finally, promoter-centered capture Hi-C from the hippocampus is included to show the potential long-range chromatin interactions across the genomic neighborhood.

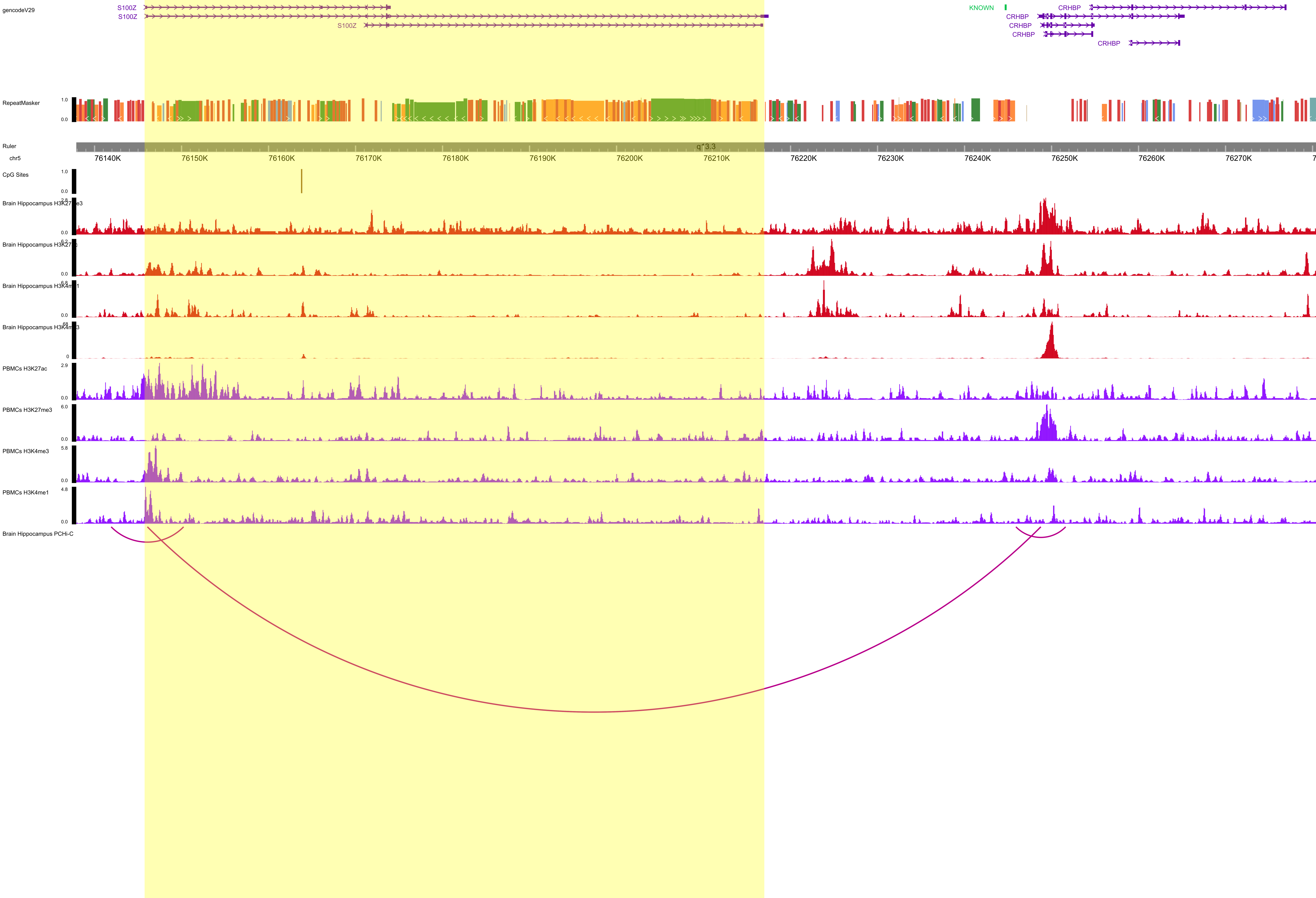

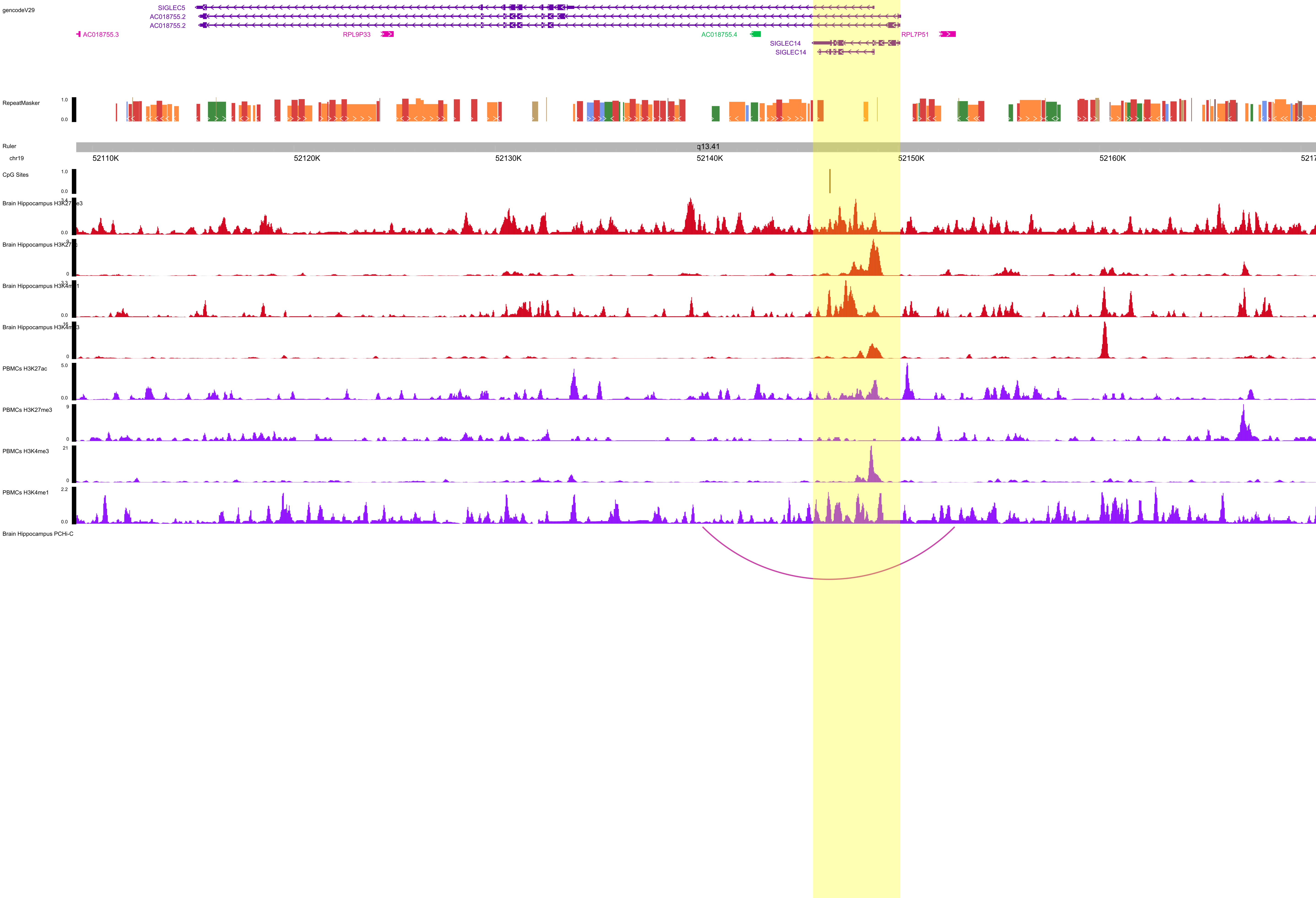

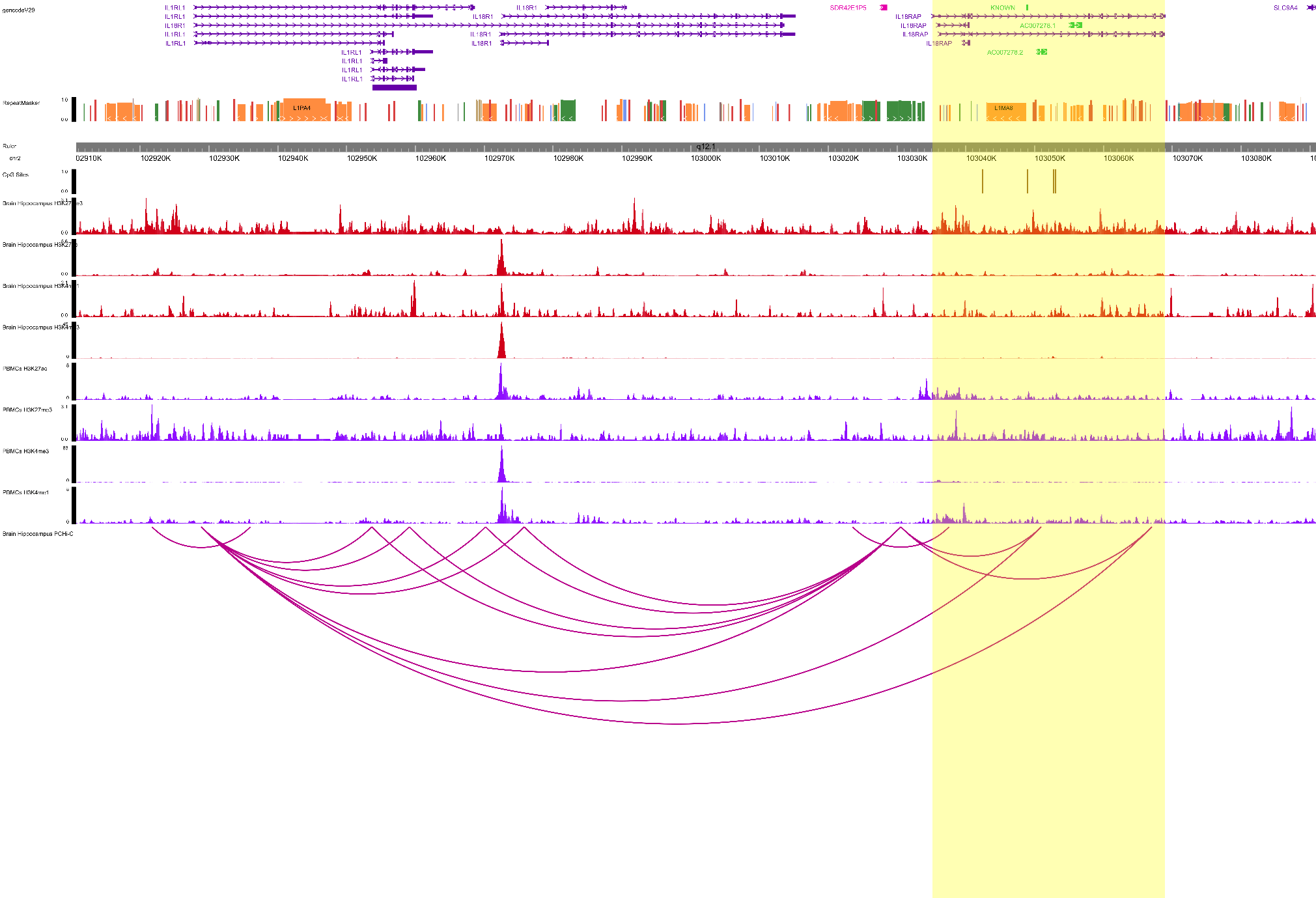

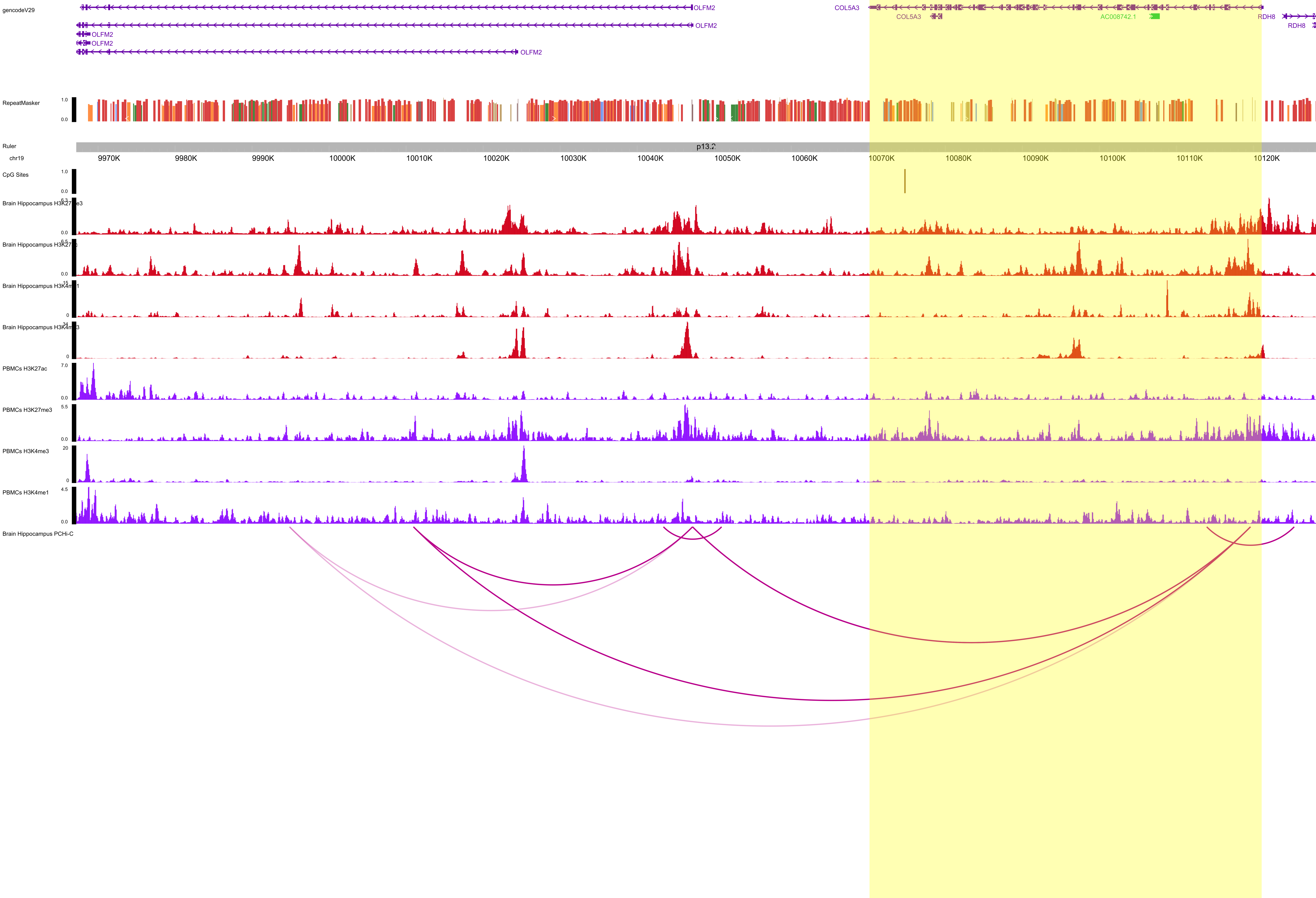

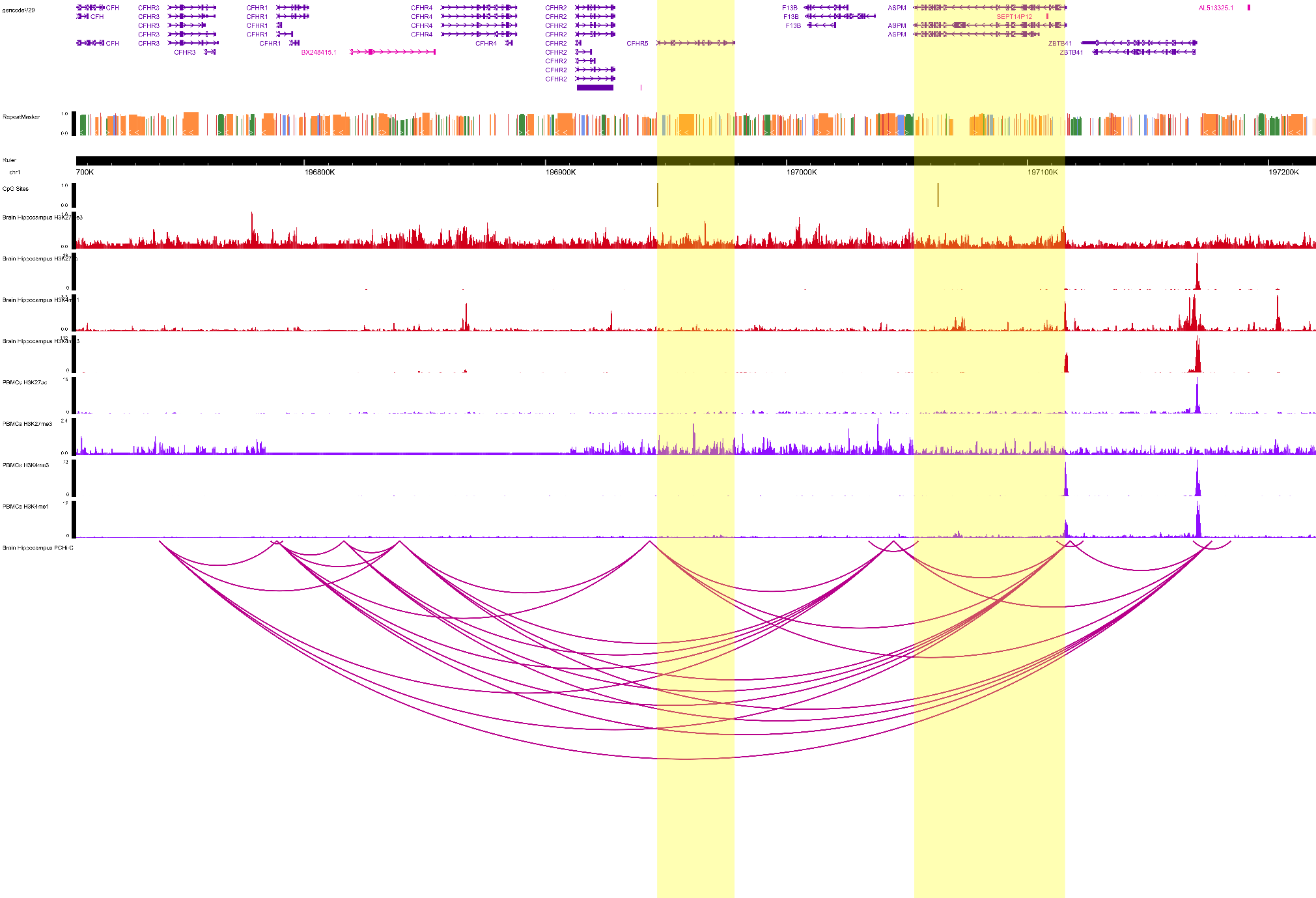

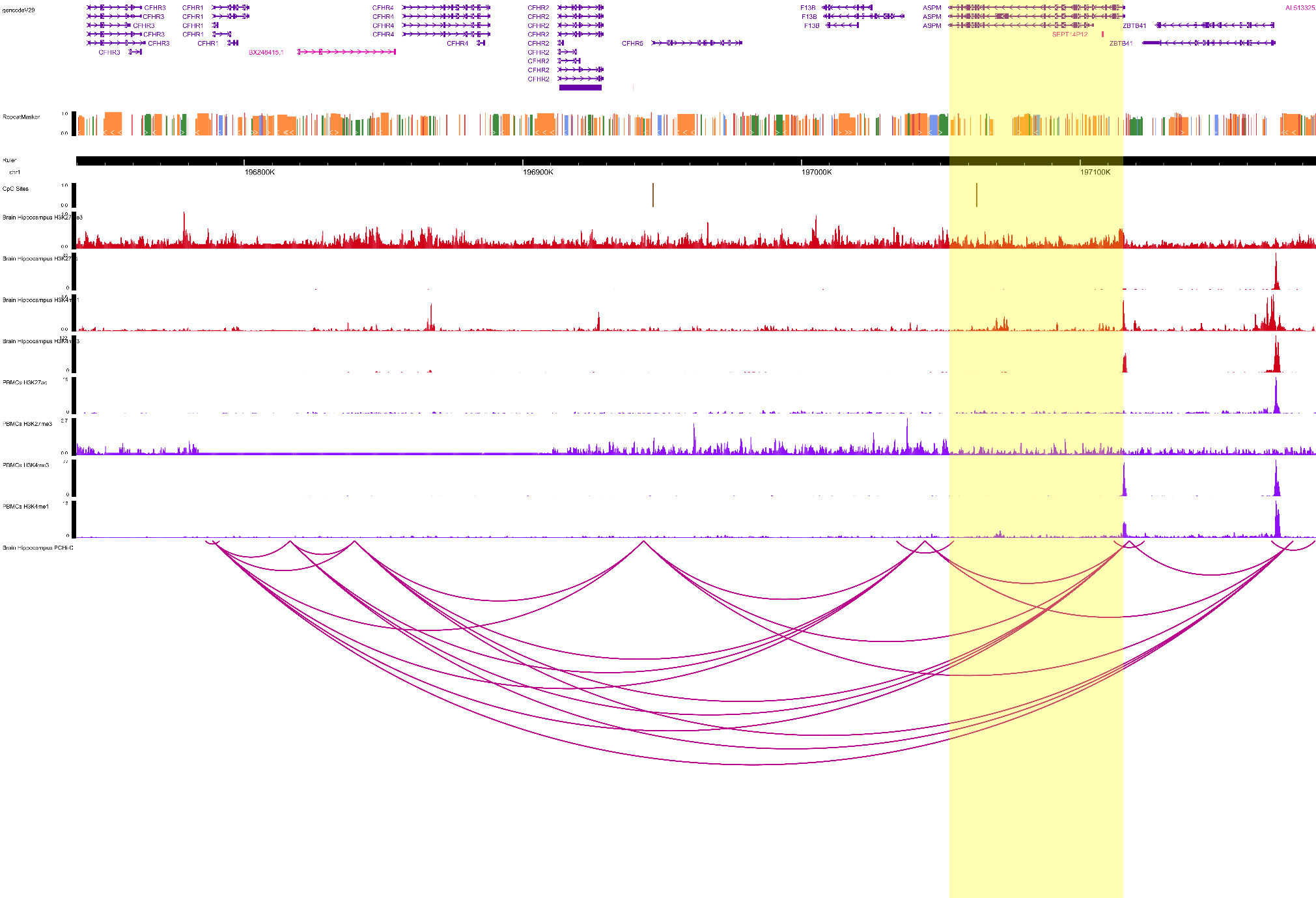

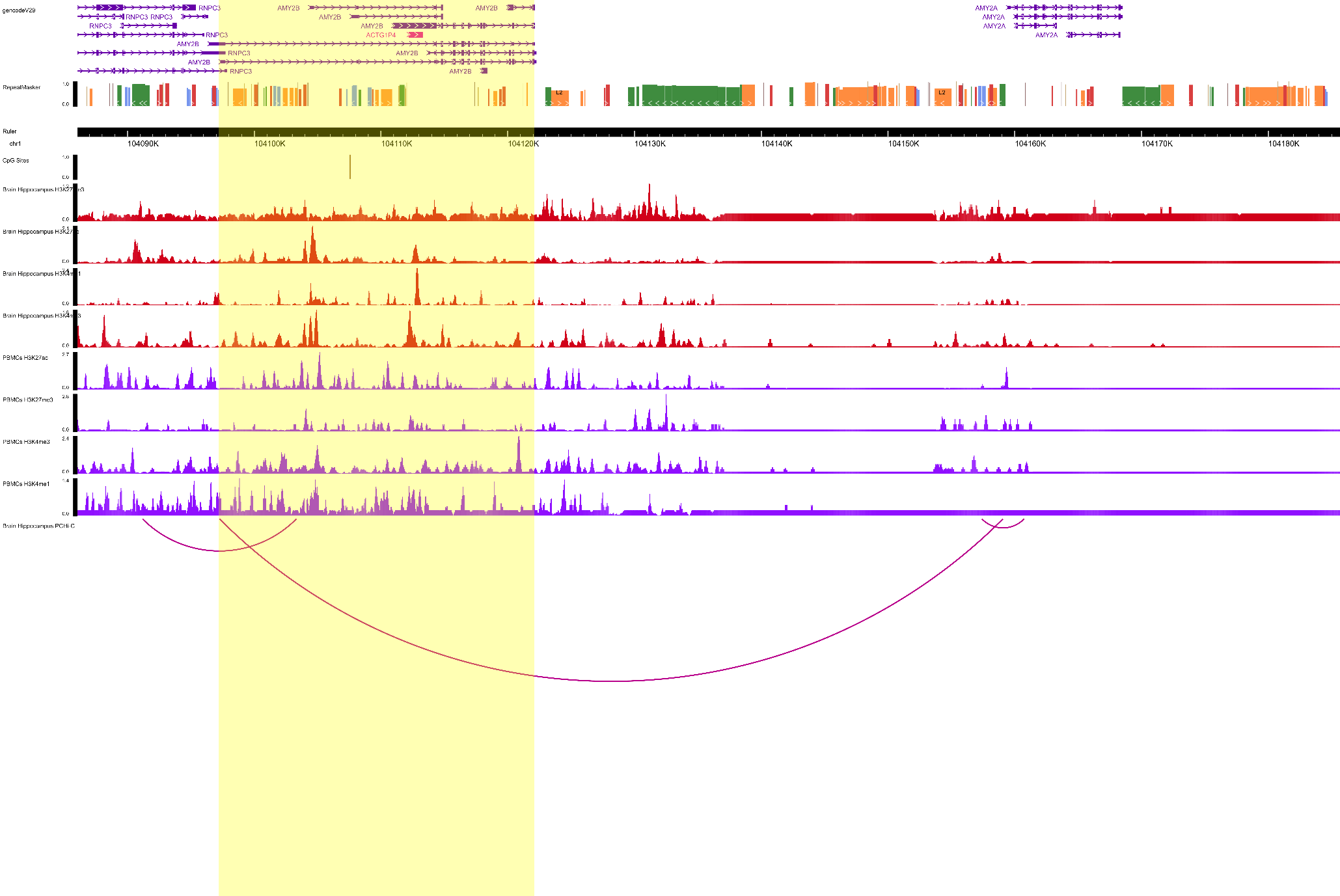

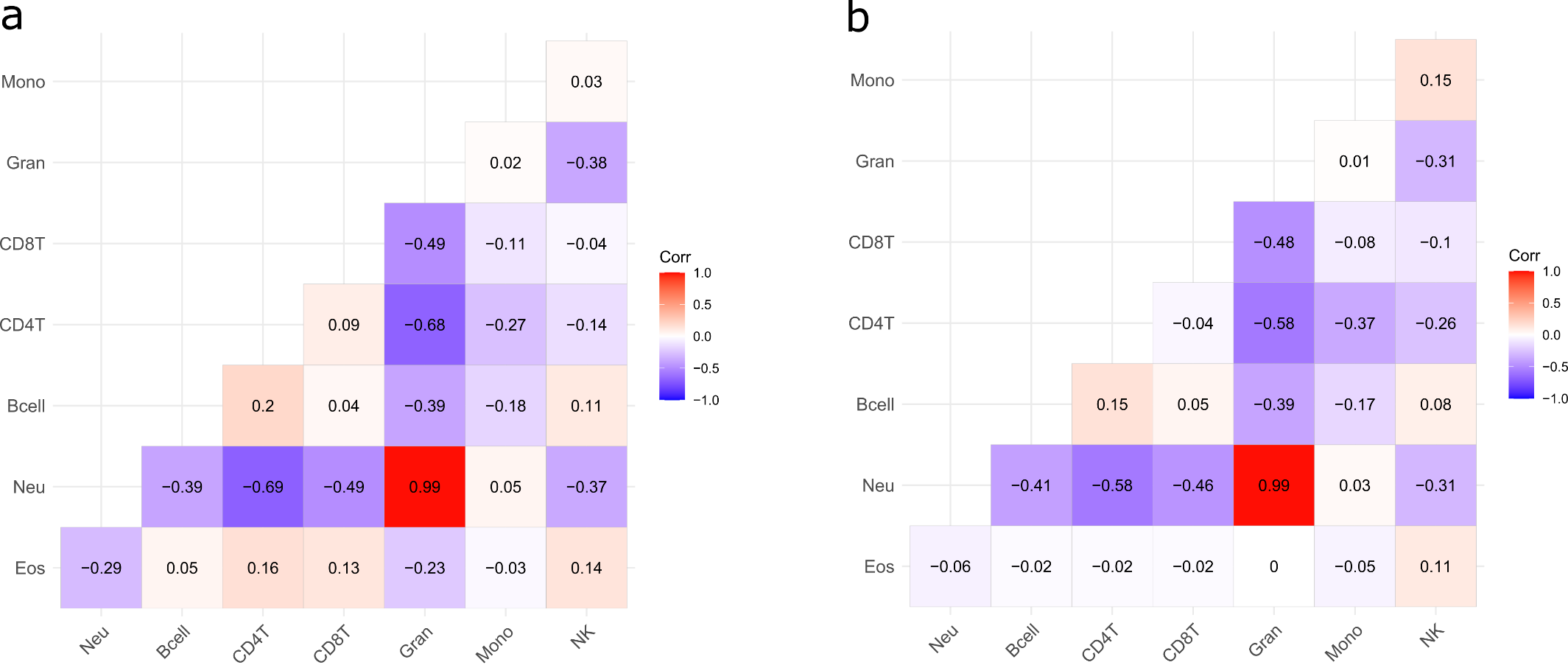

**Supplementary Figure 22. Correlation heatmaps for the white blood cell estimates available for MWAS analyses (N=774) in Set 1 (a) and Set 2 (b).** Pearson’s correlation coefficients (*r*) are provided as annotations. There were 476 and 298 individuals in Sets 1 and 2, respectively. Mono: Monocytes. Gran: Granulocytes. CD8T: CD8+ (cytotoxic) T cells. CD4T: CD4+ (cytotoxic) T cells. Bcell: B cells (lymphocytes). Neu: Neutrophils. Eos: Eoisonophils.
